# Coinfection with Respiratory Pathogens in COVID-19 in Korea

**DOI:** 10.1101/2020.12.18.20248449

**Authors:** Kyoung Ho Roh, Yu Kyung Kim, Shin-Woo Kim, Eun-Rim Kang, Yong-Jin Yang, Sun-Kyung Jung, Sun-Hwa Lee, Nackmoon Sung

**Affiliations:** Clinical Research Institute/Molecular Diagnosis Center, Seegene Medical Foundation, Seoul, Rep. of Korea; Department of Clinical Pathology, School of Medicine, Kyungpook National University Hospital, Daegu, Rep. of Korea; Division of Infectious Disease, Department of Internal Medicine, Kyungpook National University Hospital, Daegu, Rep. of Korea

**Keywords:** COVID-19, Coinfection, Respiratory Pathogens, Influenza

## Abstract

Detection of severe acute respiratory syndrome coronavirus 2 (SARS-CoV-2) in upper and lower respiratory specimens and coinfection with other respiratory pathogens in patients with coronavirus disease 2019 (COVID-19) were investigated. From the study subjects (N = 258) retrospectively enrolled when confirmed as SARS-CoV-2 positive, nasopharyngeal (NPS), oropharyngeal swabs (OPS), and sputum specimens were restored for retesting SARS-CoV-2 and detecting respiratory pathogens. Majority of the study subjects (95.7%, N = 247) were confirmed as SARS-CoV-2 positive using NPS/OPS specimens, suggesting that the upper respiratory specimen is most valuable in detecting SARS-CoV-2. Coinfection rates in COVID-19 patients (N = 258) with respiratory pathogens were 9.7% (N = 25); 8.5% (N = 22) respiratory viruses and 1.2% (N = 3) *Mycoplasma pneumoniae*, an atypical bacterium. Of the respiratory virus coinfection cases (N = 22), 20 (90.9%) were co-infected with a single respiratory virus and 2 (0.8%) (metapneumovirus/adenovirus and rhinovirus/bocavirus 1/2/3/4) with two viruses. Respiratory viruses in single viral coinfection cases with SARS-CoV-2 were as follows: non-SARS-CoV-2 coronaviruses (229E, NL63, and OC43, N = 5, 1.9%), rhinovirus (N = 4, 1.6%), metapneumovirus (N = 3, 1.2%), influenza A (N = 3, 1.2%), respiratory syncytial virus A and B (N = 3, 1.2%), and adenovirus (N = 2, 0.8%). No mixed coinfections with respiratory viruses and *M. pneumonia*e were found. In conclusion, the diagnostic value of utilizing NPS/OPS specimen is excellent, and, as the first report in Korea, coinfection with respiratory pathogens were detected at a rate of 9.7% in patients with COVID-19.

## Introduction

COVID-19, caused by the infection of SARS-CoV-2, was identified as a cluster of pneumonia cases in Wuhan, China, in December 2019, and has spread to other countries since then, resulting in 42 million cases and over 1.1 million deaths globally as of October 25, 2020 (1).

In Korea, a Chinese female from Wuhan, China, was identified as the first COVID-19 case on January 19, 2020, during the quarantine inspection at Incheon airport (2). Suspects with respiratory symptoms and/or a history of travel to other countries, including China, were asked to test for SARS-CoV-2 infection, causing exponential growth by conducting real-time PCR (RT-PCR) tests to identify COVID-19 using emergency use authorized (EUA) *in vitro* diagnostics (IVD) assays (3).

For the fast and accurate diagnosis of SARS-CoV-2 using IVD assays, the selection of appropriate types of specimens collected from patients at the right time would be an important factor (4). It has been reported that lower respiratory specimens such as bronchoalveolar lavage fluid and sputum have been recommended as the best clinical respiratory specimens for detecting SARS-CoV-2 (5, 6). Viral loads of SARS-CoV-2 were higher in nasal swabs than those in throat swabs collected from symptomatic COVID-19 patients (7). The Centers for Disease Control and Prevention recommended upper respiratory specimens as acceptable for initial diagnostic testing of SARS-CoV-2 in the types of nasopharyngeal swab (NPS), oropharyngeal swab (OPS), nasal swab, and saliva (8). Simultaneous collection and placement of NPS and OPS in a universal transport medium (U™) tube using two sets of swabs is recommended to increase sensitivity in real-time polymerase chain reaction (RT-PCR) assays in Korea, as long as the supply of flocked swabs is not limited (9). We analyzed the number of patients with COVID-19 that were confirmed as positive using NPS/OPS by measuring the detection rates of SARS-COV-2 among the patients diagnosed during the screening process in the first outbreak of the disease in Korea.

The IVD assays for detecting SARS-CoV-2 were not incorporated for testing coinfections with other respiratory pathogens at the time of screening COVID-19 suspects. Recently, coinfections in COVID-19 have been reported not to neglect infections by other respiratory pathogens in addition to SARS-CoV-2 (10-14). This suggests that simultaneous testing for coinfections between SARS-CoV-2 and other respiratory pathogens would be required to provide a better patient treatment during the COVID-19 pandemic (15).

Among the respiratory pathogens, including tuberculosis, virus and bacteria have been predominantly reported as coinfection agents in COVID-19, as seen in the previous influenza pandemic (10, 12, 16, 17). Common coinfecting viruses with SARS-CoV-2 include respiratory syncytial virus, influenza, rhinovirus/enterovirus, parainfluenza, metapneumovirus, and non-SARS-CoV-2 coronaviruses, and the coinfecting bacteria included *Mycoplasma pneumoniae, Legionella pneumophila, Chlamydia pneumoniae, Pseudomonas aeruginosa, Haemophilus influenzae*, and *Streptococcus pneumoniae* (10-13).

In this study, we simultaneously detected frequently reported respiratory viruses and atypical bacteria in clinical specimens collected from patients with COVID-19 to identify coinfection rates using RT-PCR-based commercial assays. Upper (NPS and OPS) and lower (sputum) respiratory tract specimens were employed to investigate the coinfections with respiratory pathogens when COVID-19 was confirmed as positive during the screening process.

This study reports simultaneous detection of SARS-CoV-2 and coinfection with respiratory pathogens (viruses and atypical bacteria) using commercially available RT-PCR assays among COVID-19 patients confirmed during the first outbreak in Korea.

## Materials and Methods

### Study subjects

Request forms of COVID-19 suspects submitted to Seegene Medical Foundation, a Non-Profit Independent Clinical Reference Laboratory, for detecting SARS-CoV-2 from February 9 to 23, 2020, were retrospectively reviewed. The request forms were generated and recorded by government healthcare centers, hospitals, or quarantine offices with personal and clinical information of COVID-19 suspects, such as name, address, age, birth date, sex, specimen type and collection date, and respiratory symptoms including history of visiting foreign countries, based on the guidelines of the Korea Disease Control and Prevention Agency (KDCA). Once request forms with respiratory tract specimens (NPS/OPS and/or sputum) collected from the suspects were submitted, the test results were reported within 24 h of turnaround time. Based on the guidelines for testing COVID-19 in Korea, both upper respiratory tract samples (NPS and OPS) were placed in a tube containing a U™ to increase test sensitivity (9, 18, 19).

We reviewed the request forms and enrolled study subjects who were COVID-19 positive for fever (> 37.5°C) and cough because these two respiratory symptoms were reported as the most common (10, 16). The respiratory specimens (U™ containing NPS/OPS as upper respiratory specimen and sputum as lower respiratory specimen) of the study subjects were restored from storage freezers (−70°C) and applied to extract nucleic acids to reconfirm SARS-CoV-2 and detect respiratory pathogens simultaneously.

### Specimen preparation and nucleic acid extraction

Specimens collected from the enrolled study subjects were subjected to nucleic acid extraction as described below for detecting SARS-CoV-2 and respiratory pathogens. A portion (200 µL) of the U™ containing NPS/OPS was directly applied to the nucleic acid extraction. The sputum specimens were inspected for viscosity, homogenized, and diluted with phosphate-buffered saline as recommended by the specimen treatment guidelines of the Korean Society for Laboratory Medicine (9, 18) Then, 200 µL of diluted sputum was applied for nucleic acid extraction.

Nucleic acid extraction was carried out by mixing the specimen (U™ or diluted sputum) and ready-to use reagents of a commercial kit (MagNA Pure 96 DNA and Viral NA Small Volume Kit; Roche Applied Diagnostics, Mannheim, Germany) as recommended by the manufacturer. The mixtures were placed in MagNA Pure 96 instruments (Roche Diagnostics) and processed for nucleic acid extraction according to the manufacturer’s instructions. The final elution volume of nucleic acids extracted from each specimen was approximately 100 µL. The extracted nucleic acids were stored at - 70°C and used for detecting SARS-CoV-2 and respiratory pathogens.

### Detection of SARS-CoV-2 and respiratory pathogens using RT-PCR

To detect SARS-CoV-2, a commercial RT-PCR assay (Allplex™2019-nCoV Assay, Seegene Inc., Seoul, Republic of Korea) was employed in this study. Briefly, the extracted nucleic acids (8 µL) were mixed with reagents of the commercial RT-PCR assay, such as primers and probes for specifically detecting three target genes of the virus [envelope protein (E) gene, RNA-dependent RNA polymerase (RdRP) gene, and nucleocapsid protein (N) gene] (20), exogenous internal control, and Real-time One-step Enzyme in an automated liquid handling workstation (STARlet, Seegene Inc.) to maintain accuracy and prevent human errors. After that, RT-PCR reactions were performed in a CFX96 real-time PCR cycler (Bio-Rad, Hercules, CA, USA) as recommended by the manufacturer for detecting SARS-CoV-2.

The nucleic acids extracted from the specimens of the study subjects were also applied to simultaneously detect respiratory pathogens using commercially available multiplex real-time PCR assays (Allplex™Respiratory Panel 1, 2, and 3 assay for viruses and Allplex™PneumoBacter assay for atypical bacteria, Seegene Inc.). The respiratory viruses detected in this study were influenza A and B (Flu A and Flu B), respiratory syncytial virus A and B (RSV), adenovirus (Adv), metapneumovirus (MPV), enterovirus (HEV), parainfluenza viruses 1, 2, 3, and 4 (PIV 1, 2, 3, and 4), bocavirus 1/2/3/4 (HBoV), rhinovirus (HRV), and other coronaviruses (229E, NL63, and OC43). Atypical bacteria such as *M. pneumoniae, C. pneumoniae, L. pneumophila*, and *Bordetella pertussis* were simultaneously detected with the respiratory viruses. Interpretation for positive and negative detection of respiratory pathogens by RT-PCR was determined as recommended by the manufacturer of the multiplex assays.

### Statistical Analysis

The statistical significance of the difference in detection rates of respiratory pathogens was analyzed using the Chi-square test or Fisher’s exact test. The mean value comparison of age and Ct values among the study subjects was evaluated using the Mann–Whitney test. Statistical analysis was conducted using GraphPad Prism software (Version 5.0; GraphPad Software, Inc., La Jolla, CA, USA) and GraphPad InStat software (Version 3.0). P values < 0.05 were considered statistically significant.

### Ethics statement

We retrospectively analyzed the request forms and RT-PCR assay results of the study subjects, which were exempted from informed consent. This study was approved by the Institutional Review Boards of Seegene Medical Foundation (SMF-IRB-2020-003) and Kyungpook National University Hospital (DGIRB202003001-HE002).

## Results

### Study subject enrollment

A total of 258 study subjects with fever and cough were enrolled in this study. After retrospectively reviewing the request forms and test results of suspects (N = 20,054) conducted for screening COVID-19 from February 7 to 23, 2020, we found that 404 patients (2.0%) were confirmed as SARS-CoV-2 positive based on the Allplex 2019-nCoV assay. Among the positives, patients without personal information (N = 62; 15.3%), such as age, sex, and region where they live, were excluded. Additionally, patients with no fever or cough as clinical symptoms (N = 84; 20.8%) were also excluded, and 258 subjects who were SARS-CoV-2 positive and had fever (higher than 37.5°C) and cough were enrolled.

The mean age ± standard error of the mean (SEM) of the study subjects was 48.5 ± 0.8 y (standard deviation: 15.4, 95% CI: 46.6–50.4) with a median age of 52.0 y. The detection rates of SARS-CoV-2 by the age of the study subjects were 1.2% (N = 3) for those below 20 years of age, 26.4% (N = 68) for those between 20 and 39 y, 46.1% (N = 119) for those between 40 and 59 y, and 26.4% (N = 68) for those aged > 60 y. This indicates that SARS-CoV-2 was mostly detected at the age of 40–59 y in this study.

The ratio of males and females was 41.1% (N = 106) and 58.9 % (N = 152), respectively. Residence area of the study subjects were predominantly Kyungpook Province, including Daegu city (N = 241, 93.4%), where the first outbreak of COVID-19 occurred in February, followed by Seoul (N = 6, 2.3%), Chungnarm Province (N = 5, 1.9%), and Kangwon Province (N = 3, 1.1%).

### Detection of SARS-CoV-2 by types of specimen

Study subjects (N = 258) were retested for SARS-CoV-2 using the Allplex 2019-nCoV assay. The study subjects were classified as positive for SARS-CoV-2 in both NPS/OPS and sputum (53.9%, N = 139), positive in NPS/OPS but negative in sputum (2.3%, N = 6), and negative in NPS/OPS but positive in sputum (4.3%, N = 11) (Fig. 1). Some patients (39.5%, N = 102) were SARS-CoV-2 positive in NPS/OPS, although their sputum specimens were not submitted because they were not able to expectorate (Fig. 1). Collectively, when the NPS/OPS specimen was applied without sputum, the positive rate of SARS-CoV-2 with the Allplex 2019-nCoV assay was 95.7% (N = 247) among the study subjects (N = 258), suggesting that NPS/OPS would be the most valuable specimen for molecular diagnosis for detecting SARS-CoV-2, especially in the screening of COVID-19 (Fig. 1).

**Figure 1.**
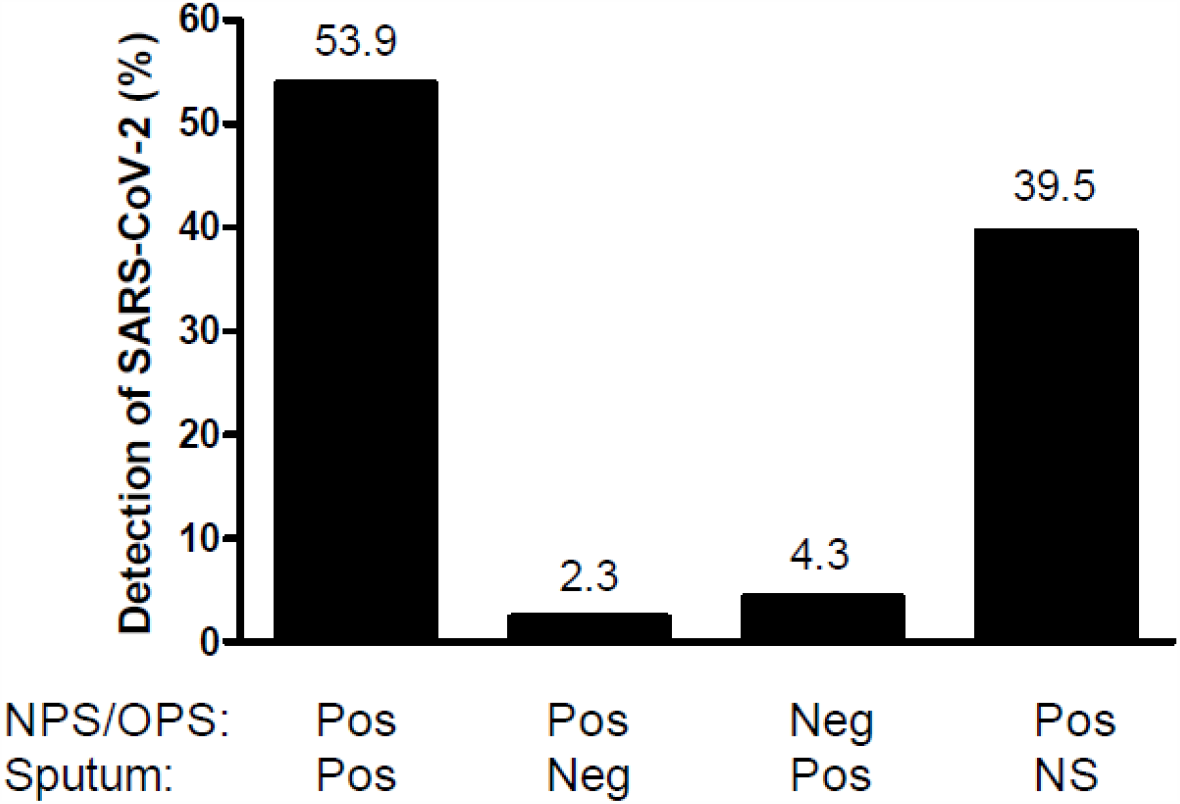
Classification of the study subjects by different types of respiratory specimen. Upper (NPS/OPS) and lower (sputum) respiratory specimens of the study subjects were retested for severe acute respiratory syndrome coronavirus 2 (SARS-CoV-2) using Allplex 2019-nCoV Assay. The study subjects (N=258) were confirmed to be positive for SARS-CoV-2 in both NPS/OPS and sputum at 53.9% (N = 139), positive in NPS/OPS but negative in sputum at 2.3% (N = 6), negative in NPS/OPS but positive in sputum at 4.3% (N = 11). Some (39.5%, N = 102) were positive in NPS/OPS when sputum samples were not submitted. NPS: Nasopharyngeal swab, OPS: Oropharyngeal swab, NS: Specimen were not submitted for COVID-19 (Coronavirus disease 2019) testing, Pos: Positive, Neg: Negative

The study subjects were confirmed to be positive for SARS-CoV-2 when all three target genes in the Allplex 2019-nCoV assay were positive, as recommended by the Korea COVID-19 Diagnosis Testing Management Committee (18). Figure 2 shows the mean cycle threshold (Ct) values of target genes for SARS-CoV-2 in the Allplex 2019-nCoV assay. The mean Ct values for all study subjects with NPS/OPS (N = 247) were 20.9 ± 0.3 (95% CI: 20.2–21.6) for the E gene, 22.3 ± 0.3 (95% CI: 21.6–22.9) for the RdRP gene, and 23.6 ± 0.3 (95% CI: 22.8–24.3) for the N gene. In sputum (N = 150), the mean Ct values for E, RdRP, and N genes were 21.9 ± 0.4 (95% CI: 21.1–22.9), 23.3 ± 0.4 (95% CI: 22.4–24.2), and 25.1 ± 0.4 (95% CI: 24.3–26.0), respectively. The Ct value for the E gene came first, RdRP gene second, and N gene third in the Allplex 2019-nCoV assay applied in this study, and the differences (delta) of the mean Ct values for E, RdRP, and N genes between NPS/OPS and sputum were 1.0, 1.0, and 1.5, respectively. The mean Ct values for the E gene and RdRP gene detected in NPS/OPS were not significantly different from those in sputum (P ≥ 0.072), but the mean Ct values for the N gene between NPS/OPS and sputum were significantly different (P = 0.0055) (Fig. 2 A). The N gene in sputum was detected significantly later than that in NPS/OPS when the Allplex 2019-nCoV assay was applied in the same patient.

**Figure 2.**
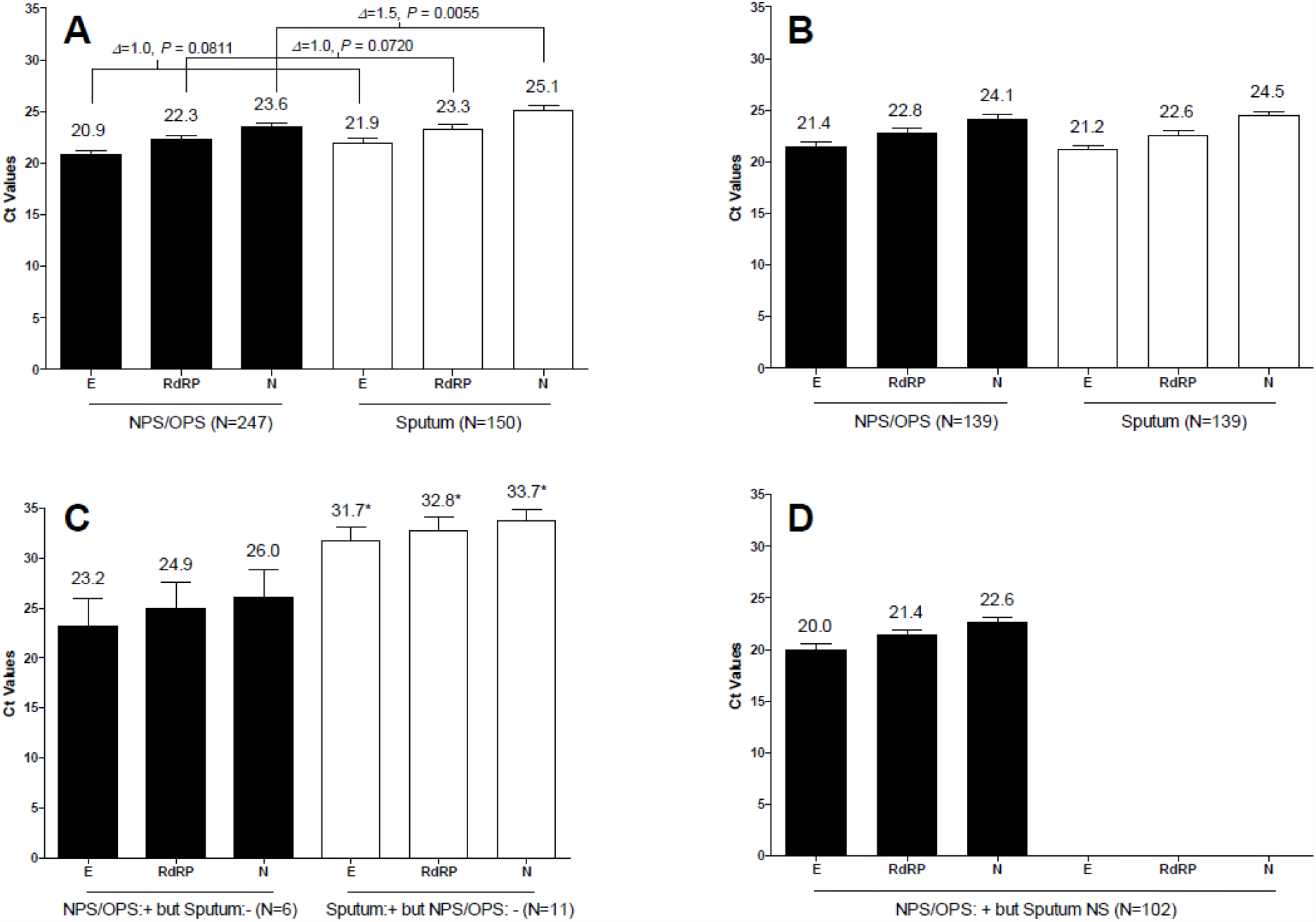
Ct values of E gene, RdRP gene, and E gene in Allplex 2019-nCoV assay for SARS-CoV-2 extracted from NPS/OPS and sputum collected from the study group (N=258). A. Ct values of E gene, RdRP gene and N gene detected in NPS/OPS (N=247) and sputum (N=150) among all of the study subjects (N=258). B. Ct values among the study subjects (N=139) classified as positive for SARS-CoV-2 in both NPS/OPS and sputum. C. Ct values among the study subjects (N=6) classified as positive for SARS-CoV-2 in NPS/OPS but negative in sputum (NPS/OPS + but sputum -) and the subjects (N=11) as negative for SARS-CoV-2 in NPS/OPS but positive in sputum (NPS/OPS – but sputum +). D. Ct values among the study subjects (N=102) classified as positive for SARS-CoV-2 in NPS/OPS even though sputum specimen were not submitted. Ct: Cycle threshold, SARS-CoV-2: Severe acute respiratory syndrome coronavirus 2, NPS: Nasopharyngeal swab, OPS: Oropharyngeal swab, E: Envelope protein gene, RdRP: RNA-dependent RNA polymerase gene, and N: Nucleocapsid protein gene * indicates significantly different; differences in Ct values of E gene, RdRP gene, and N gene between SARS-CoV-2 positive in NPS/OPS but negative in sputum (N = 6) and negative in NPS/OPS but positive in sputum (N = 11) were 8.5, 7.9, and 7.4, respectively, which were significantly different (P ≤ 0.0145).

For the study subjects who were SARS-CoV-2 positive in both NPS/OPS and sputum (53.9%, N = 139) (Fig. 1), the mean Ct values for the E, RdRP, and N genes in NPS/OPS were not significantly different (21.4 ± 0.5, 22.8 ± 0.5, and 24.1 ± 0.5, respectively) from those (21.2 ± 0.4, 22.6 ± 0.4, and 24.5 ± 0.4, respectively) in sputum (P < 0.0001) (Fig. 2 B). However, as shown in Figure 2 C, the mean Ct values (23.2 ± 2.8, 24.9 ± 2.6, and 26.0 ± 2.8, respectively) for the E, RdRP, and N genes in the study subjects who were SARS-CoV-2 positive in NPS/OPS but negative in sputum (2.3%, N = 6) were significantly lower than those (31.7 ± 1.3, 22.6 ± 1.3, and 24.5 ± 1.2, respectively) who were SARS-CoV-2 positive in sputum but negative in NPS/OPS (4.3%, N = 11) (P ≤ 0.0145). The Ct values and viral load of SARS-CoV-2 are inversely proportional; therefore, the viral loads of SARS-CoV-2 in sputum were significantly lower than those in NPS/OPS. The viral loads of SARS-CoV-2 may influence the difference in the detection rates of the virus between NPS/OPS and sputum.

The patients in whom NPS/OPS specimens were positive although sputum was not submitted (39.5%, N = 102) (Fig. 1), the mean Ct values (20.0 ± 0.5, 21.4 ± 0.5, and 22.6 ± 0.5, respectively) for E, RdRP, and N genes in NPS/OPS specimen (Fig. 2 D) were significantly lower than those of other study subjects confirmed as SARS-CoV-2 positive with the same specimen as shown in Figure 2 B and C (P ≤ 0.0407). Interestingly, although the patients were not able to expectorate sputum, the viral loads of SARS-CoV-2 in the NPS/OPS specimen were higher than that of patients who were able to expectorate sputum.

### Coinfection with Respiratory Pathogens in COVID-19

Coinfection rates with respiratory viruses and atypical bacteria among the study subjects (N = 258) were 8.5% (N = 22) and 1.2% (N = 3), respectively, which were significantly different (P = 0.0001) (Table 1). Of the respiratory virus coinfection cases (N = 22), 20 cases (90.9%) of SARS-CoV-2 positive were coinfected with a single respiratory virus and two cases (0.8%) (MPV/Adv and HRV/HBoV) were coinfected with two viruses. Respiratory viruses in single viral coinfection cases with SARS-CoV-2 were as follows: non-SARS-CoV-2 coronaviruses [N = 5, 1.9%; 229E (N = 3), NL63 (N = 1), and OC43 (N = 1)], HRV (N = 4, 1.6%), MPV (N = 3, 1.2%), Flu A (N = 3, 1.2%), RSV (N = 3, 1.2%), and Adv (N = 2, 0.8%).

**Table 1.** Study subjects (N=258) coinfected with respiratory pathogens in COVID-19.

| Patients Coinfected with Respiratory Pathogens |  | No of Patients Coinfected (%) | M:F | Mean Age (SEM) | Region of Residency (No. of Patients) | Classification of the Coinfections by Types of Specimen (N=25) |  |  |
| --- | --- | --- | --- | --- | --- | --- | --- | --- |
|  |  |  |  |  |  | No. (%) of SARS-CoV-2 Positive in: |  |  |
|  |  |  |  |  |  | NPS/OPS | Sputum | Both |
| Virus (N=22, 8.5%) | Flu A | 3 (1.2) | 1:2 | 30.3 (14.7) | KP (3) | 2* | - | 1 |
|  | RSV | 3 (1.2) | 0:3 | 47.0 (15.5) | KP (3) | - | 2 | 1 |
|  | Adv | 2 (0.8) | 2:0 | 29.0 (8.0) | KP (1), KG (1) | - | 1 | 1 |
|  | MPV | 3 (1.2) | 1:2 | 40.3 (8.0) | KP (3) | - | - | 3 |
|  | HRV | 4 (1.6) | 1:3 | 43.8 (7.7) | KP (3), Seoul (1) | 2* | - | 2 |
|  | Non-SARS-CoV-2 Coronavirus | 5 (1.9) | 3:2 | 39.8 (6.9) | KP (5) | - | 1 | 4 |
|  | MPV/Adv | 1 (0.4) | 0:1 | 53 | KP (1) | 1 | - | - |
|  | HRV/HBoV | 1 (0.4) | 0:1 | 25 | KP (1) | - | - | 1 |
| Bacteria (N=3, 1.2%) | *M. pneumoniae* | 3 (1.2) | 2:1 | 32.0 (6.0) | KP (3) | - | - | 3 |
| Total (N=25, 9.7%) |  | 25 | 10:15 | 38.4 (3.3) |  | 5 (20.0) | 4 (16.0) | 16 (64.0) |

COVID-19: Coronavirus disease 2019
Flu A; Influenza A, RSV; Respiratory syncytial virus, Adv; Adenovirus, MPV; Metapneumovirus, HRV; Rhinovirus, Non-SARS-CoV-2 Coronavirus (Coronavirus 229E, NL63, and OC43), *M. pneumoniae*; *Mycoplasma pneumoniae*, KP; Kyungpook Province, and KG; Kyunggi Province. SEM: Standard Error of Mean
\* NPS/OPS specimen were only submitted without sputum.

Among the respiratory virus coinfections (N = 22), 59.1% (N = 13) were detected in both NPS/OPS and sputum. Flu A, HRV, and MPV/Adv were detected only in NPS/OPS, and RSV, Adv, and non-SARS-CoV-2 coronaviruses were detected only in sputum (Table 1). Notably, four cases of coinfection (two patients with Flu A and two with HRV) provided only NPS/OPS specimens without sputum (Table 1). The data in this study suggest that both types of specimens should be employed for detecting coinfections with respiratory pathogens in COVID-19.

All cases of respiratory bacterial coinfection (N = 3; male:female = 2:1) were *M. pneumoniae* detected in both NPS/OPS and sputum specimens without any other atypical bacteria detected in this study (Table 1). Additionally, no mixed coinfection of viruses and bacteria among the study subjects was observed.

The ratio of males to females in the coinfection cases was 4:6 (Table 1), but no difference in coinfection by sex was observed (P > 0.05), suggesting that both men and women confirmed that COVID-19 would be susceptible to respiratory pathogens at the same level.

The mean age of coinfection cases (N = 25, 9.7%) with respiratory pathogens (both virus and bacteria) was 38.4 ± 3.3 y, which was significantly lesser than the mean age (49.6 ± 1.0) of patients without coinfections (P = 0.0009). The mean ages for viral (N = 22) and bacteria (N = 3) coinfection cases were 39.2 ± 3.6 y and 32.0 ± 6.0 y, respectively, which were not statistically significant (P = 0.4841) (Table 1). Most of the coinfection cases resided in Kyungpook Province, except two cases (coinfections of Adv and HRV in Kyunggi Province and Seoul, respectively) (Table 1).

Figure 3 shows the age distribution of coinfection cases (N = 25) with respiratory pathogens among the study subjects (N = 258). The rates of coinfections in the age groups of < 20 years old, between 20 and 39, between 40 and 59, and > 60 were 0.4% (N = 1), 4.3% (N = 11), 4.3% (N = 11), and 0.8% (N = 2), respectively. Coinfections with Flu A were detected at ages below 20 y (N = 1) and between 40 and 59 y (N = 2). At age > 60 y, both RSV and other coronaviruses were coinfected. Two cases (0.8%) of double viral coinfections were found at ages between 20 and 39 y (HRV/HBoV) and between 40 and 59 y (MPV/Adv), respectively. All other coinfection cases (7.0%, N = 18) with virus and *M. pneumoniae* were detected between 20 and 59 years of age.

**Figure 3.**
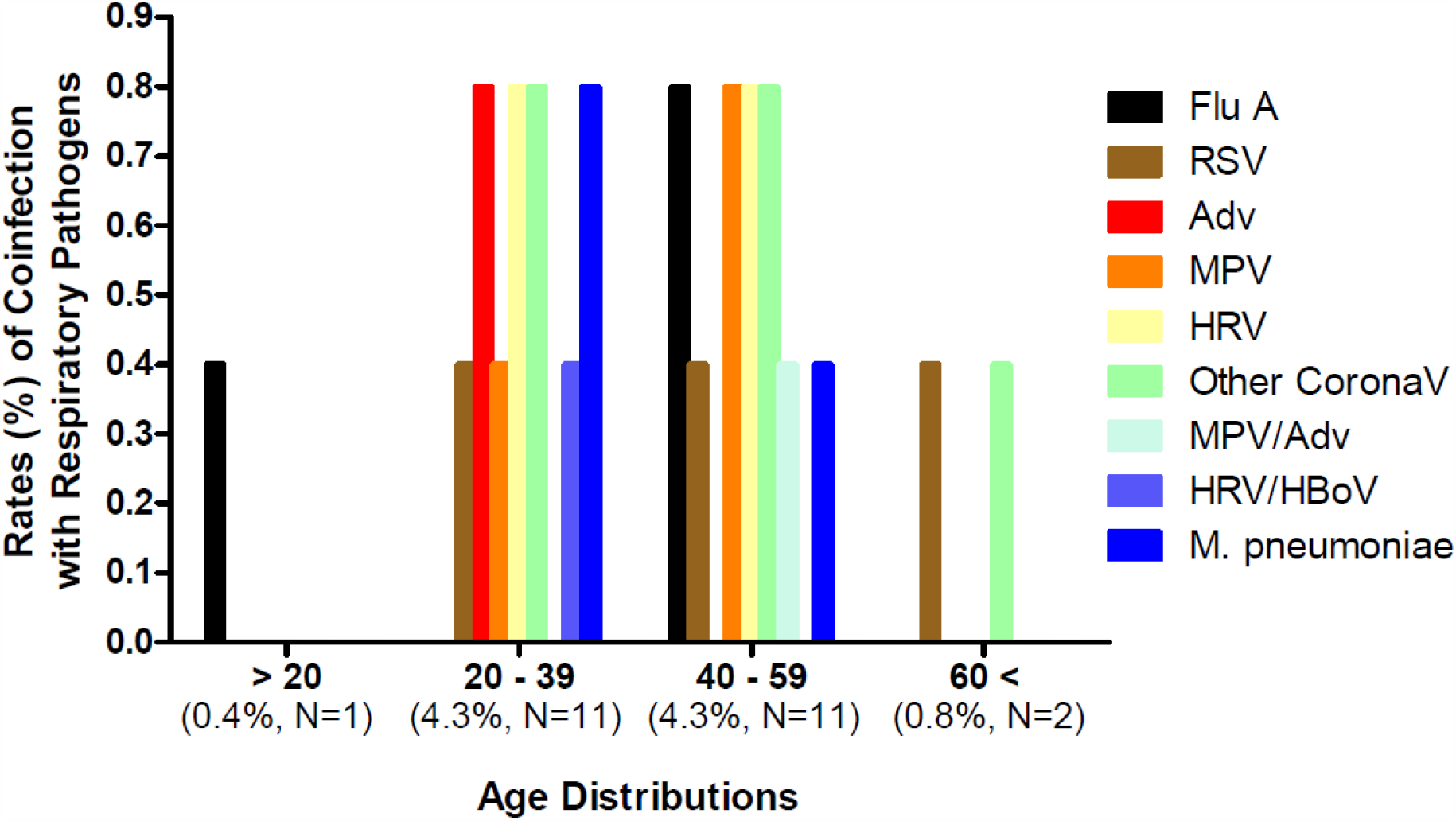
Distribution of respiratory pathogens by different ages in the coinfection cases (N = 25) in COVID-19 among the study subjects (N = 258). The numbers of coinfection cases based on the age distributions were 1 (0.4%), 11 (4.3%), 11 (4.3%), and 2 (0.8%) for ages below 20, between 20 and 39, between 40 and 59, and above 60, respectively. COVID-19: Coronavirus disease 2019 Flu A; Influenza A, RSV; Respiratory syncytial virus, Adv; Adenovirus, MPV; Metapneumovirus, HRV; Rhinovirus, Other CoronaV; Coronavirus such as 229E, NL63, and OC43, and *M. pneumoniae*; *Mycoplasma pneumoniae*,

Ct values of target genes (E, RdRP, and N) specific to SARS-CoV-2 in coinfection cases with respiratory pathogens were not significantly different from those in non-coinfection cases in the patients of COVID-19 when applying the Allplex 2019-nCoV assay, regardless of the type of respiratory specimen (P ≥ 0.2771).

## Discussion

In Korea, both NPS and OPS were placed in a U™ tube after collection from suspects of COVID-19 for screening (9). The U™ tubes were transported for COVID-19 testing to the laboratory immediately after collection, with or without sputum samples. No other upper or lower respiratory tract samples were transported and tested. Among the suspects (N = 20,054) tested from February 9 to 23, 2020, during the first outbreak in Korea, the positive rate of SRAS-CoV-2 was 2.0% (N = 404) using the RT-PCR assay, which was lower than the finding (9.5%; N = 116) in a previous study, when tested for patients in the USA (N = 1,217) enrolled from March 3 to 25, 2020 (21). From the positive (N = 404), patients (N=258) with clinical symptoms (fever and cough) were enrolled as study subjects for this study.

Among the study subjects (N = 258), 95.7% (N = 247) were confirmed to be positive for SARS-CoV-2 in the RT-PCR assay by sampling NPS/OPS, regardless of the collection of sputum (Fig. 1). Additionally, based on a comparison of the mean Ct values for the genes specific to SARS-CoV-2, higher viral loads of SARS-CoV-2 were observed in NPS/OPS than in sputum (Fig. 2), which would support the excellent detection rates of SARS-CoV-2 in NPS/OPS (Fig. 1). This suggests that NPS/OPS would be the most valuable for diagnosing SARS-CoV-2 in molecular diagnosis assays, and it would not be necessary to enforce sampling sputum specimens, especially when the patients were not able to expectorate. However, in previous studies, sputum specimens were considered a better choice than nasal samples for the molecular detection of SARS-CoV-2. Pan et al. (6) measured the viral loads of SARS-CoV-2 from two COVID-19 patients at the early stage of onset of symptoms in throat, sputum, urine, and stool. They found that the viral loads were higher in sputum samples than in throat samples, especially in the early days after onset. Wang et al. (5) compared the detection rates of SARS-CoV-2 among multiple types of specimens collected from patients with COVID-19, bronchoalveolar lavage fluid showed 93% positive rate as the highest, followed by sputum (72%), nasal swab (63%), and pharyngeal swabs (32%). Collectively, based on the literature published, for detecting SARS-CoV-2 in the patients, the best clinical specimen recommended was bronchoalveolar lavage fluid, and sputum was the second with strong recommendation, whereas pharyngeal swabs were in moderate (22, 23). In contrast, our data support that pharyngeal swab samples would be better than sputum in detecting SARS-CoV-2 among the study subjects who underwent COVID-19 screening. A consistent finding with the current study was also observed in a case study of two Korean patients, reporting that viral loads of upper respiratory specimens (placing both NPS and OPS in the same tube) were detected as similar to or sometimes higher than those in sputum using RT-PCR (24).

Coinfection rate in patients with COVID-19 (N = 258) with respiratory pathogens was 9.7% (N = 25) in the current study (Table 1 and Figure 3): 8.5% (N = 22) of viral and 1.2% (N = 3) of atypical bacterial coinfections. The rate of viral coinfection has been previously reported to range from 2.0% – 19.8% in different countries (21, 25-27). The coinfection rate with respiratory viruses was 2.0% (N = 39) among 1,996 patients with COVID-19 hospitalized in New York City (25). Tested at a hospital in Shenzhen, China, six (3.2%) of 92 COVID-19 patients hospitalized were viral coinfection cases (26). Another study conducted in Wuhan, China, reported that among patients with COVID-19 (N = 104), 5.8% (N = 6) had coinfections with non-SARS-CoV-2 coronaviruses (2.9%, N = 3), influenza A (2.9%, N = 3), and rhinovirus (1.9%, N = 2) (27). In the study by Kim et al. (21) with patients in California, USA, 19.8% (N = 23) of SARS-CoV-2 positive patients (N = 116) were positive for other respiratory pathogens. As the first report in Korea representing the suspect group of COVID-19, especially from the first outbreak area in February 2020, 8.5% of viral coinfections were found in this study, which is comparable with previous reports.

Among the coinfections (N=25) by respiratory virus (N = 22) and the atypical bacteria *M. pneumoniae* (N = 3), most cases were detected in both the NPS/OPS and sputum specimens (64.0%, N=16), but some were in either NPS/OPS (20.0%, N=5) or sputum (16.0%, N=4) (Table 1). This suggests that both upper and lower respiratory tract specimens would be recommended to monitor the coinfection with respiratory pathogens in COVID-19. This is consistent with a previous report by Zhu et al. (14), who suggested that for detecting coinfections with respiratory pathogens in COVID-19, both upper and lower respiratory tract specimens should be collected and considered to test while diagnosing and treating COVID-19.

Based on a meta-analysis by Lansbury et al. (12) with the previously reported studies regarding types of co-infected respiratory viruses in patients with SARS-CoV-2, the most common co-infecting virus was RSV, followed by Flu A, HRV, PIV, and other coronaviruses. A study in China, which was not included in the meta-analysis, consistently reported that RSV was most commonly detected among the coinfection viruses in COVID-19 patients diagnosed between January 19 and February 26, 2020 (15). However, in the USA, Kim et al. (21) and Richardson et al. (25) detected rhinovirus/enterovirus as the most common coinfection agent in their study, followed by RSV among the study subjects enrolled in March 2020. In this study with Korean study subjects enrolled in February 2020, the most common respiratory virus as the coinfection agent was HRV [1.9 %, N = 5; one double viral coinfection with HBoV (HRV/HBoV) and four single viral coinfection cases in SARS-CoV 2 positive] (Table 1). Five cases of single viral coinfections by non-SARS-CoV-2 coronaviruses (three cases of 229E, one case of NL63, and one case of OC43) were also observed (Table 1).

SARS-CoV-2 and influenza coinfection was relatively rare in three patients (1.2%) among the study subjects (N = 258) (Table 1). This finding is also consistent with previous studies, such as 0.54% in Turkey (13), 0.9% (21) and 2.4% (25) in the USA, and from 1.2%–4.3% in China (14, 15, 27, 28).

Among the atypical bacteria tested in this study, only *M. pneumoniae* was detected at the level of 1.2% (N = 3) of the study subjects (N = 258) (Table 1). In some other studies, however, *C. pneumoniae*, another atypical bacterium, was nominated as the coinfection agent in patients with COVID-19 in addition to *M. pneumoniae* (14, 15, 25). The coinfection rates in the above studies for *M. pneumoniae* and *C. pneumoniae* ranged from 1.6%–4.8% and 2.5%–5.2%, respectively. Lansbury et al. (12) emphasized, in their meta-analysis study, that *M. pneumoniae* was the most common bacteria detected in patients with COVID-19 having coinfections with respiratory bacterial pathogens, followed by *P. aeruginosa, H. influenza, Klebsiella pneumoniae*, and *Chlamydophila* spp., suggesting that in addition to coinfection by atypical bacteria, other respiratory bacteria were also the candidates as co-infecting agents in COVID-19.

Unlike Kim et al. (21), who reported no difference in age between coinfection and SARS-CoV-2 only (non-coinfection), the mean ages between them were significantly different in this study (38.4 ± 3.3 y vs. 49.6 ± 1.0 y, respectively; P = 0.0009). This suggested that young ages were more susceptible to coinfection with respiratory pathogens in this study.

The viral loads of SARS-CoV-2 between the patients with coinfection and SARS-CoV-2 only infection were not significantly different as Ct values for the target genes (E, RdRP, and N) were statistically the same between them when the Allplex 2019-nCoV assay was applied (P > 0.05). This suggests that the viral load of SARS-CoV-2 is not responsible to cause coinfection with respiratory pathogens.

The current study has a few limitations. The effect of coinfection on the treatment outcomes of COVID-19 patients enrolled in this study is unavailable. The study subjects in this study were retrospectively enrolled during the process of screening COVID-19 in the region of the first outbreak in Korea. Thus, the scope of this study was to investigate the coinfection rates of respiratory pathogens among patients with COVID-19, not hospitalized, but diagnosed as positive during the screening process for SARS-CoV-2. Additionally, the results of the study cannot represent the coinfection rates for the winter when respiratory pathogens such as influenza are dominant. The study subjects were only enrolled in February; therefore, investigating the coinfection rates with respiratory pathogens would be interesting during the whole winter season, especially from November to March in Korea.

In summary, the upper respiratory tract specimen (NPS/UPS) provided excellent detection of SARS-CoV-2 without lower respiratory specimens (sputum) in the RT-PCR assay. However, detection of coinfections with respiratory pathogens in COVID-19 required both upper and lower respiratory specimens. Coinfection rates in patients with COVID-19 (N = 258) with respiratory pathogens were 9.7% (N = 25) with mostly virus (8.5%, N = 22) and an atypical bacterium, *M. pneumoniae* (1.2%, N = 3). The most common respiratory virus detected as a coinfecting agent was HRV, followed by non-SARS-CoV-2 coronaviruses, HRV, MPV, Flu A, RSV, and Adv. As a conclusion, simultaneous detection of respiratory pathogens and SARS-CoV-2 by molecular diagnosis assays would be necessary for identifying the causative agents causing coinfection, especially during the COVID-19 pandemic.

## Data Availability

Data would be available upon request. 
Please contact the corresponding author, Nackmoon Sung, PhD via eamil.

## Acknowledgements

The authors appreciate Prof. Doosu Jeon at Busan National University Yangsan Hospital for his critical review and comments. This research was funded in part by the KOICA IBS program (2018-0119). The current study was supported by the Korea Disease Control and Prevention Agency (KDCA) by COVID-19 Testing Service contracted to the Seegene Medical Foundation.

